# Novel autoantibodies help diagnose anti-SSA antibody negative Sjögren’s disease and predict abnormal labial salivary gland pathology

**DOI:** 10.1101/2023.08.29.23294775

**Authors:** Maxwell Parker, Zihao Zheng, Michael Lasarev, Roxana A. Alexandridis, Michael A. Newton, Miriam A. Shelef, Sara S. McCoy

## Abstract

**Objectives:** Sj□gren’s disease (SjD) diagnosis requires either positive anti-SSA antibodies or a labial salivary gland biopsy with a positive focus score (FS). One-third of SjD patients lack anti-SSA antibodies (SSA-), requiring a positive FS for diagnosis. Our objective was to identify novel autoantibodies to diagnose ‘seronegative’ SjD.

**Methods:** IgG binding to a high density whole human peptidome array was quantified using sera from SSA- SjD cases and matched non-autoimmune controls. We identified the highest bound peptides using empirical Bayesian statistical filters, which we confirmed in an independent cohort comprising SSA- SjD (n=76), sicca controls without autoimmunity (n=75), and autoimmune controls (SjD features but not meeting SjD criteria; n=41). In this external validation, we used non-parametric methods for peptide abundance and controlled false discovery rate in group comparisons. For predictive modeling, we used logistic regression, model selection methods, and cross-validation to identify clinical and peptide variables that predict SSA- SjD and FS positivity.

**Results:** IgG against a peptide from D-aminoacyl-tRNA deacylase (DTD2) was bound more in SSA- SjD than sicca controls (p=.004) and more than combined controls (sicca and autoimmune controls combined; p=0.003). IgG against peptides from retroelement silencing factor-1 (RESF1) and DTD2, were bound more in FS-positive than FS-negative participants (p=.010; p=0.012). A predictive model incorporating clinical variables showed good discrimination between SjD versus control (AUC 74%) and between FS-positive versus FS-negative (AUC 72%).

**Conclusion:** We present novel autoantibodies in SSA- SjD that have good predictive value for SSA- SjD and FS-positivity.

**KEY MESSAGES:**

- <u>What is already known on this topic</u> - Seronegative (anti-SSA antibody negative [SSA-]) Sjögren’s disease (SjD) requires a labial salivary gland biopsy for diagnosis, which is challenging to obtain and interpret.
- <u>What this study adds</u> - We identified novel autoantibodies in SSA- SjD that, when combined with readily available clinical variables, provide good predictive ability to discriminate 1) SSA- SjD from control participants and 2) abnormal salivary gland biopsies from normal salivary gland biopsies.
- <u>How this study might affect research, practice or policy</u> - This study provides novel diagnostic antibodies addressing the critical need for improvement of SSA- SjD diagnostic tools.

## INTRODUCTION

Sj□gren’s disease (SjD) is an autoimmune exocrinopathy with characteristic focal lymphocytic infiltrate of salivary glands (SGs) that results in symptoms of oral and ocular dryness. Although patients most commonly experience exocrine gland-related symptoms, over 40% of individuals have extra-glandular systemic organ involvement [1].

The diagnosis of SjD is challenging. Dryness is common, present in up to 65% of the general population [2]; however, SjD has a prevalence of less than 1% [3]. Unlike dryness attributed to many other causes, dryness from SjD is caused by autoimmunity. Detecting autoimmunity, and thus diagnosing SjD, requires either a positive anti-SSA antibody test or a labial salivary gland biopsy with a focus score (FS) ≥ 1 (i.e. ≥1 foci [50 mononuclear cells] per 4 mm^2^ of tissue).

Accurate diagnostic testing is critical because patients with SjD need to be followed longitudinally for extraglandular organ involvement and appropriate targeted therapeutic intervention. Anti-SSA antibody is present in 40-68% of SjD patients [1]. Thus, about a third of SjD patients are anti-SSA antibody negative (seronegative or SSA-). This ‘seronegative’ patient population requires a labial salivary gland biopsy for diagnosis. A specialist is required to perform the biopsy and pathologists experienced in FS calculation must interpret results. The latter requirement is often overlooked, but re-evaluation of salivary gland biopsies by expert pathologists results in a diagnostic revision in over half of cases [4]. Moreover, a labial salivary gland biopsy is invasive with a rare risk of focal numbness. Understandably patients can be reluctant to undergo this procedure. Thus, procuring and interpreting labial salivary gland biopsies are limiting steps toward a timely diagnosis of SjD [5].

To address the challenges associated with labial SG biopsies, a major need in the SjD community is new biomarkers to diagnose SSA- SjD. Ideally biomarkers have high sensitivity and specificity and use specimens that are readily available (e.g., blood, tears, or saliva). In this study, we identified novel autoantibodies in seronegative SjD sera using a whole human peptidome array and confirmed our results with ELISA.

## METHODS

### Population

For the human peptidome array and validation ELISAs, we used sera from eight anti-SSA antibody negative (SSA-) SjD participants meeting SjD American College of Rheumatology (ACR)/ European Alliance of Associations for Rheumatology (EULAR) 2016 criteria [6] and 8 age- and sex-matched controls without autoimmune or inflammatory disease (Table 1) as previously described from the University of Wisconsin rheumatology Biorepository (IRB# 2015-0156) [7]. Each autoimmune disease participant had an age- and sex-matched control with some control participants serving as a control for more than one autoimmune disease participant.

**Table 1.** Demographics of participants.

| Peptidome Array and Internal Validation (n=16) |  |  |  |  |
| --- | --- | --- | --- | --- |
|  | SSA- SjD<br>(n=8) | Control<br>(n=8) |  |  |
| Age mean (SD) | 58 (12) | 59 (10) |  |  |
| Female n (%) | 8 (100) | 8 (100) |  |  |
| White n (%) | 8 (100) | 8 (100) |  |  |
| Hispanic n (%) | 0 | 0 |  |  |
| External Validation (n=192) |  |  |  |  |
|  | SSA- SjD<br>(n=76) | Autoimmune<br>Control<br>(n=41) | Sicca<br>Control<br>(n=75) | p-value |
| Age mean (SD) | 55 (12) | 55 (12) | 55 (12) | 1 |
| Female Sex n (%)* | 65 (86) | 33 (87) | 64 (85) | 0.93 |
| Race n (%) |  |  |  | 0.18 |
| White/Hispanic | 45 (59) | 26 (63) | 41 (55) |  |
| Asian | 31 (41) | 54 (37) | 30 (40) |  |
| African | 0 | 0 | 4 (5) |  |
| Clinical Metrics n (%) |  |  |  |  |
| OSS ≥ 5 | 56 (76) | 16 (39) | 20 (27) | <0.0001 |
| Schirmer's ≤ 5 mm/5min | 39 (53) | 10 (26) | 22 (32) | 0.005 |
| UWS ≤ 0.5 mL/5min | 52 (68) | 16 (39) | 31 (41) | 0.001 |
| Lab metrics |  |  |  |  |
| Platelet k/μL | 248.2 (65.5) | 257.9 (72.6) | 269.1 (85.9) | 0.2 |
| ANA positive ≥1:320 | 22 (29) | 12 (29) | 0 | <0.0001 |
| IgG mg/dL mean (SD) | 1343.2<br>(764.8) | 1357.10<br>(703.4) | 1058.3<br>(342.5) | 0.008 |
| RF positive n (%) | 31 (41) | 27 (66) | 0 | <0.0001 |
| anti-SSB positive n (%) | 31 (41) | 27 (66) | 0 | <0.0001 |
| Histopathology |  |  |  |  |
| Focus score ≥ 1 | 76 (100) | 9 (22) | 0 | <0.0001 |

OSS=ocular staining score; UWS=unstimulated whole salivary flow; focus score = the number of foci (50 mononuclear cells) per 4 mm<sup>2</sup> of tissue.

For external validation, we used samples from the Sjögren’s International Collaborative Clinical Alliance (SICCA) registry and biorepository, a multisite international registry housed at the University of California, San Francisco. Participants were enrolled in the SICCA registry if they had i) a known diagnosis of SjD, ii) salivary gland enlargement, iii) repeated dental caries without risk factors, or iv) abnormal serology (anti-SSA or anti-SSB antibody, antinuclear antibody [ANA], or rheumatoid factor [RF]). Further registry details can be found at https://siccaonline.ucsf.edu or as described in prior publications [6, 8, 9]. In addition to IRB approval obtained for each SICCA clinical research site, and all foreign institutions housing these sites having Federal wide Assurance, we obtained IRB approval from the UW health sciences IRB (IRB # 2021-0945) to perform the analyses presented in this paper.

All SSA- SjD participants met the 2016 ACR/EULAR criteria. We compared SSA- SjD participants (n=76) to sicca-controls (n=75) and autoimmune-controls (n=38; Table 1). Sicca-controls had symptoms or signs of dryness but lacked autoimmunity (ANA < 1:320, negative RF, negative anti-SSA antibodies, and FS <1 on labial salivary gland biopsy). Autoimmune controls had autoimmune features (ANA ≥ 1:320, positive RF, or FS ≥1 on labial salivary gland biopsy) but did not meet the 2016 ACR/EULAR criteria for SjD.

### Patient and public involvement

Patients/public were not involved in the design, conduct, or reporting of the manuscript.

### Whole peptidome array and statistical analysis

To evaluate autoantibody reactivity, identify common features of antigens, and better understand SjD, we used a whole human peptidome array (Roche NimbleGen, Madison WI), whose general technology we previously validated [10]. The peptidome array was composed of over 5.3 million overlapping 16 amino acid peptides tiled at 2 amino acid intervals across the human proteome. Although methods to analyze large data sets on gene expression exist, antibody binding to peptide arrays have different sampling features and require unique techniques to differentiate signal from noise. In order to account for signal and noise characteristics of the peptide array, we used a large-scale testing tool, MixTwice [11], and r-value [12] to prioritize peptides for differential signal intensity between two groups. These empirical Bayesian tools enable ranking and calibration via local false discovery rate (locFDR) in low signal/noise settings by accounting for shared attributes of peptide-specific sampling distributions.

We assigned peptides a locFDR for sensitive filtering, combined with data on binding affinity, protein context, and peptide sequence. We defined a nearest neighbor peptide as on the same protein and at the immediate neighboring position. The nearest neighbor locFDR of a peptide is the averaged locFDR of its nearest neighbor peptide(s). We used a combination of r-value <0.01, locFDR <0.01, and nearest neighbor locFDR <0.05 on peptides transformed by empirical cumulative distribution function and found 469 peptides bound more in seronegative SjD than controls and 431 peptides bound less in seronegative SjD than controls (Figure 1).

**Figure 1.**
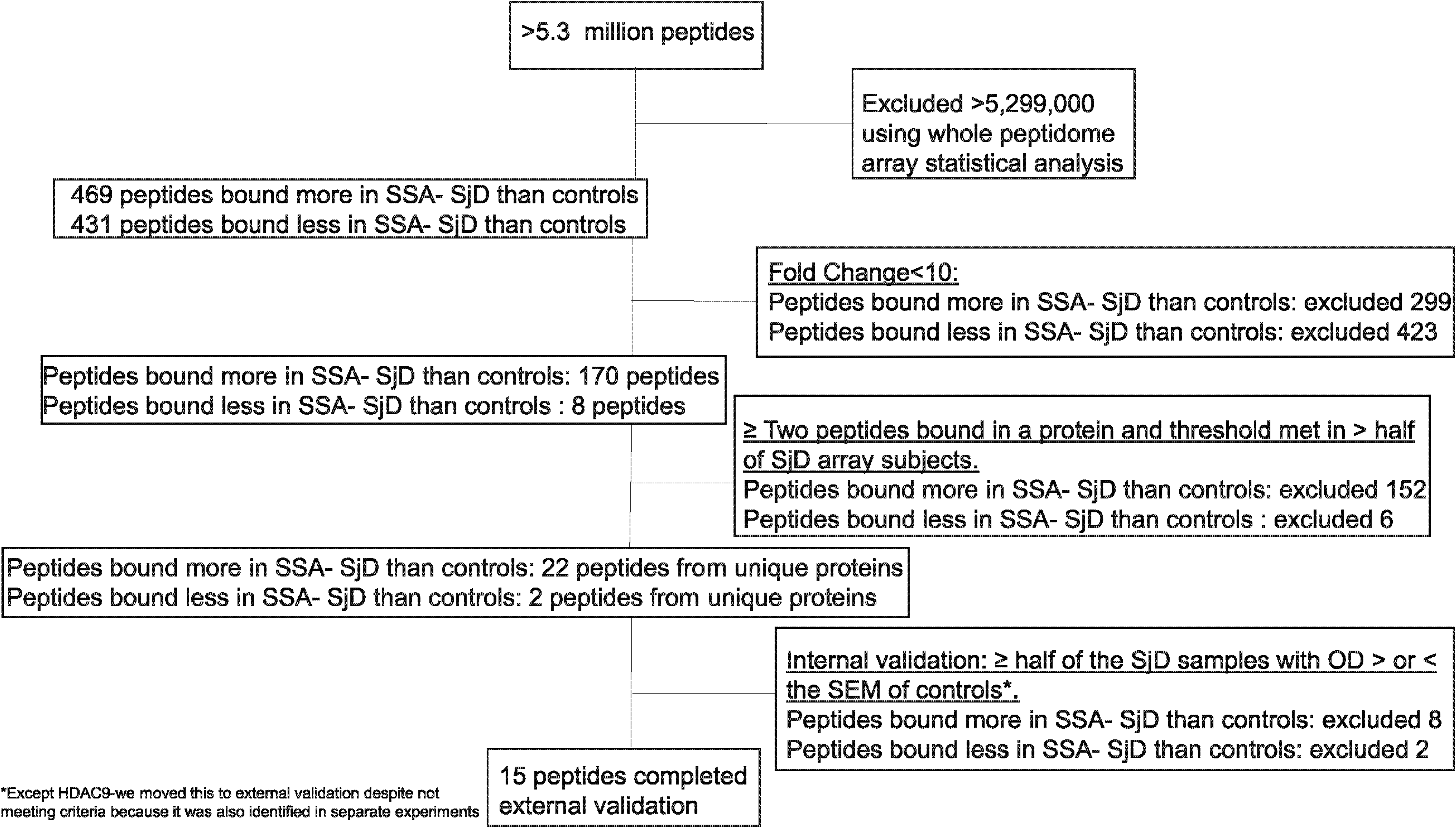
Consort flow diagram demonstrating selection of top peptides for ELISA confirmation. Starting with over 5.3 million peptides on the human peptidome array, we reduced our peptides of interest to 469 peptides bound more in SSA- SjD than controls and 431 peptides bound less in SSA- SjD than controls with our whole peptidome array analysis. This was narrowed to 22 peptides bound more in SSA- SjD than controls and 2 peptides bound less in SSA- SjD than controls by narrowing to those peptides with a fold increase in SSA- SjD versus control of at least 10, requiring at least two significant peptides bound in the same protein, and at least half of participant bound more than a threshold (threshold defined as mean plus one standard deviation of all peptide signals on the array). 15 candidate peptides were ultimately selected for external validation after removing peptides where less than half of the SjD values were beyond the standard error of the mean (SEM) of control participants.

### Selection of peptides for ELISA validation of whole peptidome array

We narrowed our peptides by requiring a fold increase in SSA- SjD versus controls of at least 10, at least two significant peptides to be bound in the same protein, and at least half of SjD participants bound more than a predefined threshold (mean plus one standard deviation of all peptide signals on the array). The resulting 24 peptides (Figure 1) were synthesized and used in ELISA to validate array findings with sera from the same participants.

### Selection of peptides from the internal ELISA validation of the array for external validation

Among peptides bound more by SSA- SjD than control IgG on the array validation ELISA, we selected peptides for external validation according to the following criteria: (1) average IgG binding at an optical density (OD) of ≥ 0.05 for SSA- SjD participants and (2) IgG binding of at least half of SSA- SjD participants with an OD greater than the standard error of the mean (SEM) of the control participants (Figure 1). None of the peptides that were bound less by SSA- SjD than control IgG met criteria to proceed to external validation.

### ELISA

The peptide ELISA was optimized for serum concentration, peptide concentration, and incubation duration and performed as described in the Supplemental Methods. Resulting OD values were adjusted by normalizing to the following controls: 1) blank wells, 2) wells with peptide coating but no sera, 3) wells with sera but no peptide coating, and 4) two positive controls on each plate used to normalize for plate-to-plate variation.

### Statistical analysis of ELISA results

Functional annotations and protein domains were generated using NIH DAVID 6.8 online tool to determine function (biological, cellular, and molecular processes) and protein domains (InterPro, SMART, UP_KW_Domain) of top peptides (website: https://david-d.ncifcrf.gov/) [13, 14]. Motif analysis was performed with Meme Suite [15] and PROSITE [16–18] was used to identify proteins containing the identified motifs.

Mann-Whitney, Kruskal-Wallis and Wilcoxon rank-sum tests (2-sided) were used to compare adjusted optical density of each peptide among groups of interest (Graphpad Prism software [Graphpad software, La Jolla, CA; R v4.2.2]). Two-group comparisons used one-sided p-values, the direction being confirmed from the initial array findings and internal validation. The Benjamini-Hochberg method was used to form adjusted p-values (q-values) that are adjusted for false discovery rate (FDR) within each block of tests on the (15) peptides entering external validation.

To build predictive models incorporating clinical variables, we used adaptive lasso for clinical variable selection (predicting SjD vs. sicca control and FS positive vs. negative) (Jmp Pro 17, Cary, NC). Of 21 clinical variables (Supplemental Table A), the top six identified by adaptive lasso regression included ocular staining score ≥ 5, platelet count, IgG, ANA ≥ 1:320, RF, and unstimulated whole salivary flow. Because ocular staining scores are not readily available to most clinicians, we included platelet count, IgG, ANA, RF, and unstimulated whole salivary flow into predictive model calculations that incorporate the new peptides entering external validation.

Separate logistic regression models were created to predict odds of SjD or positive FS as a function of adjusted OD for the peptides and clinical variables identified from the adaptive lasso. Graphical exploration of continuous features suggested some could benefit from transformation (log or square-root) prior to inclusion in models. Continuous features (or transformations thereof) were initially modeled using restricted cubic splines (3 knots) to allow for potential non-linear associations. Performance was quantified with Receiver Operator Characteristic (ROC) curve (*C*-statistic) and further adjusted for model optimism [19]. Final model construction and validation was performed using R (v4.2.2) [20] and the associated rms package [21]. Nagelkerke’s *R*^2^ measure [22] were used to determine optimism adjusted values. Absolute reduction in the area under the ROC curve (AUC) or in *R*^2^_N_ (versus the full model with all relevant [transformed] predictors) is given for each single-term deletion. Additionally, we used random-subsampling (i.e., Monte Carlo cross validation) to check the capacity of novel peptide binding data to improve outcome prediction beyond the use of clinical variables alone [23, 24]; we used 10,000 random splits and an 80/20 training test ratio, though results were relatively insensitive to that ratio. Within each training set, we used marginal prescreening and stepwise model selection to obtain separate logistic regressions using clinical variables only or clinical variables and peptide variables, and we compared prediction accuracy on the test sets via differences in areas under the ROC curves.

The prevalence of positive FS among patients referred for minor salivary gland biopsy (prevalence of 0.173 [95% CI: 0.113–0.254]) [25], together with positive and negative likelihood ratios (at various cut-points) was used to compute positive and negative predictive values. Confidence intervals for the P(N)PV were developed from the separate confidence intervals for the prevalence and likelihood ratios [26].

## RESULTS

### Whole human peptidome array analysis

Of >5.3 million peptides, our analysis yielded 469 peptides bound more by SSA- SjD sera than controls and 431 peptides bound less by SSA- SjD sera than controls. We identified four motifs from the peptides bound more in SSA- SjD than control participants (Figure 2A). Of these four motifs, two had hits on PROSITE to proteins relevant in SjD. Motif [HYA]-G-[YW]-G- [QG]-[ADT]-[NG]-[DTA]-[AT]-[SND]-[SYK] is found in heterogeneous nuclear ribonucleoprotein (hn RNP), A-kinase anchor protein 8-like, and serine protease 55. Motif [MP]-[GA]-F-[RP]-[GD]- [NLK]-[PD]-G-[NQK]-[FD]-[VG] is found in complement c1q tumor necrosis factor-related protein (CTRP)2 and collagenα6(IV) chain. Of the four motifs from peptides bound less in SSA- SjD than controls (Figure 2B), none could be matched on PROSITE to <100 known proteins.

**Figure 2.**
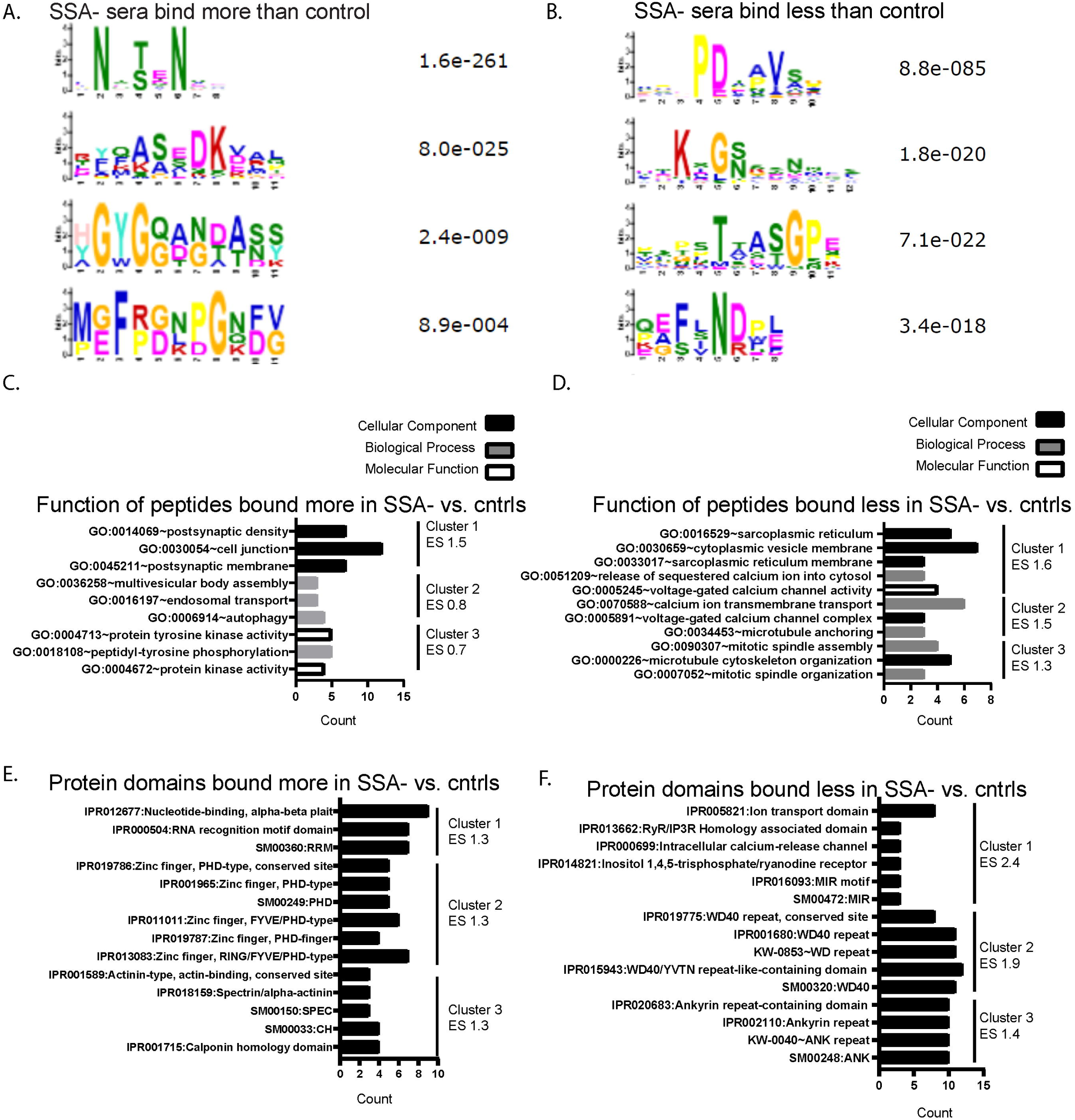
Peptide motifs, gene ontology, and functional analysis of proteins for which peptides were bound by IgG more or less in SSA- SjD than non-autoimmune controls. A) Peptide motifs bound more by SSA- SjD IgG than control IgG (n=8 participants, n=469 peptides); B) Peptide motifs bound less by SSA- SjD IgG than control IgG (n=8 participants, n=431 peptides); C) Gene ontology of peptides bound more by SSA- SjD IgG than control IgG; D) Gene ontology of peptides bound less by SSA- SjD IgG than control IgG; E) Functional protein domain binding analysis of peptides bound more by SSA- SjD IgG than healthy control sera; F) Functional protein domain binding analysis of peptides bound less by SSA- SjD IgG than control IgG. ES=enrichment score.

Among peptides bound more by SSA- SjD IgG, GO showed a top enriched cluster of post synaptic/cell junction (Figure 2C; Supplemental Table B). Among peptides bound less with SSA- SjD IgG, the top enriched GO cluster was sarcoplasmic reticulum (Figure 2D; Supplemental Table B). Top protein domains identified from peptides bound more by SSA- SjD IgG include RNA binding, zinc finger, and alpha actinin (Figure 2E; Supplemental Table C). The top protein domains from peptides bound less by SSA- SjD IgG include Ca2+ channel signaling, WD-40 repeats, and ankyrin repeats (Figure 2F; Supplemental Table C).

### Antibodies to D-aminoacyl-tRNA deacylase 2 and retroelement silencing factor 1 are higher in SSA- SjD participants than control participants

We validated our top candidate array peptides (n=24) with ELISA using the same participant sera that was used for the array (‘internal validation’). Based on the results of our internal validation (Figure 3 and Supplemental Figure A), we selected 15 peptides for external ELISA validation using different participant sera. Dot plots of external validation findings are shown in Supplemental Figure B. Using 2-sided Wilcoxon rank-sum tests, we found IgG binding to peptides from D-aminoacyl-tRNA deacylase 2 (DTD2) had an estimated 64% chance of an adjusted OD higher for a SSA- SjD than a sicca control participant (95% confidence interval [CI]: 54-72%; p=0.004; Figure 4A). We found IgG binding to peptides from retroelement silencing factor 1 (RESF1) had an estimated 59% chance of an adjusted OD higher for a SSA- SjD participant than sicca control (95% CI: 50-68%; p=0.047; Figure 4A).

**Figure 3.**
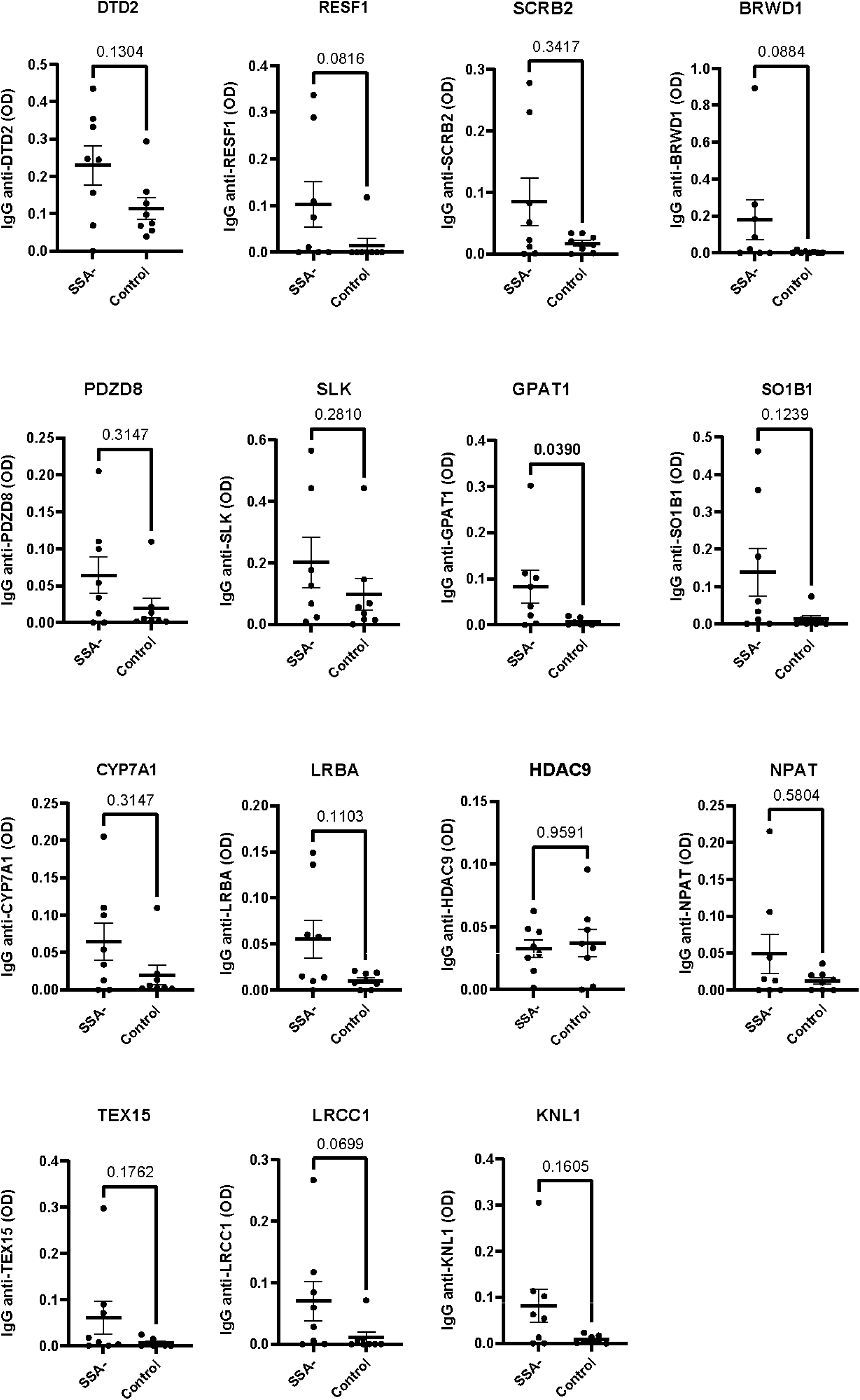
Internal validation of peptides identified from the array as bound more by SSA- SjD sera than control sera. ELISA results of IgG binding from SSA- SjD sera vs. control sera to peptides from different proteins. The participants used for these ELISAs were the same as those used on the array (n=8 SSA- SjD participants age, sex, race matched to n=8 control participants). P values reported in each panel were determined by Mann-Whitney test.

**Figure 4.**
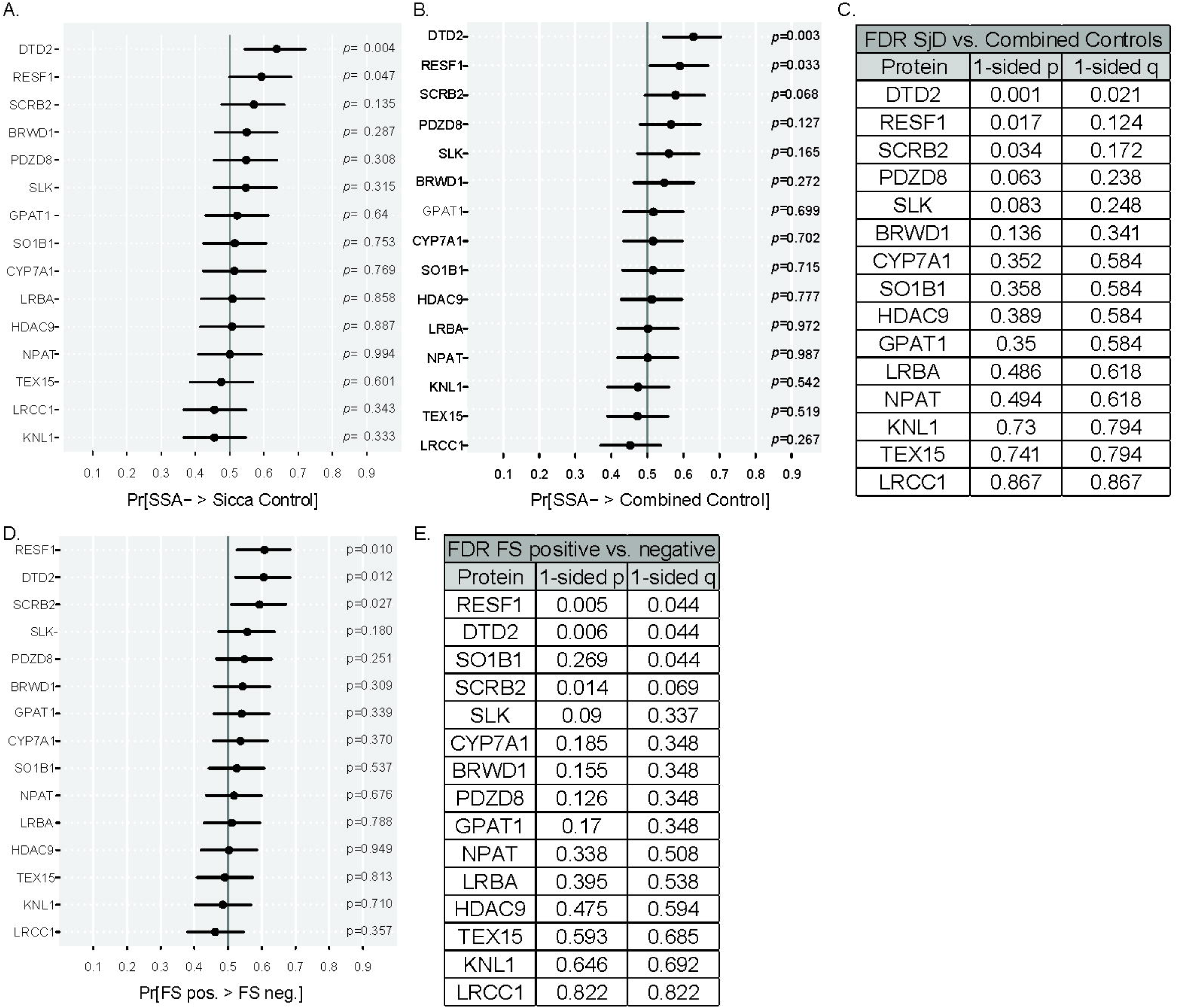
IgG from SSA- SjD and FS positive participants binds peptides from DTD2 and RESF1 more than control IgG. IgG from SSA- SjD participants bind peptides from DTD2 and RESF1 more than control IgG. IgG from FS positive participants bind peptides from RESF1, DTD2, and SCRB2 more than FS negative IgG. A) Area under the ROC curve (AUC) of the adjusted optical density of peptide groups between SSA- SjD (n=76) and sicca controls (n=75); B) AUC of the adjusted optical density of peptide groups between SSA- SjD (n=76) and combined sicca and autoimmune controls (n=116); C) One-sided Wilcoxon rank-sum test with Benjamini-Hochberg correction (q-value) for SjD vs. combined control participants; D) AUC comparing distributions of adjusted optical density of peptide groups comparing between FS positive vs. negative biopsies (n=85 FS positive and n=107 FS negative). The forest plot shows the degree of IgG binding to the peptide of interest differed between focus score positive and focus score negative comparisons; E) One-sided Wilcoxon rank-sum test with q-value of binding from FS positive vs. FS negative participants.

Next, we compared antibody binding to our top candidate peptides in SSA- SjD to a combination of sicca and autoimmune controls (combined controls). As above, peptides from DTD2 and RESF1 were bound more by SSA- SjD than combined controls IgG (p=.003 and p=0.033, respectively; Figure 4B). IgG binding to DTD2 had a 63% chance of observing a higher adjusted OD in SSA- SjD participants than combined controls (95% CI: 54-70%). Binding to RESF1 had a 59% chance of being higher in SSA- SjD participants than combined controls (95% CI: 51-67%). Recognizing the directional changes from the array data, we performed a one-sided Wilcoxon rank-sum test with Benjamini-Hochberg correction (q-value) to control the false discovery rate (Figure 4C). Binding to peptides from DTD2 survived at 5% (q=0.021).

### Antibodies to RESF1, DTD2, and SCRB2 are higher in labial salivary gland biopsies with a FS ≥ 1 than biopsies with FS < 1

Because a surrogate marker for a positive or negative labial salivary gland biopsy is a significant clinical need, we evaluated whether autoantibody binding to the 15 peptides differed between participants who had a positive biopsy (FS ≥ 1) compared to a negative FS on biopsy (FS < 1). We found that IgG from SSA- SjD participants bound peptides from RESF1, DTD2, and SCRB2 more than sera from combined control participants (p=0.010, p=0.012, p=0.027, respectively; Figure 4D). IgG to RESF1 and DTD2 both had an estimated 61% chance that adjusted OD would be higher for a positive than a negative FS (95% CI: 53-68% and 52-68%, respectively). IgG to SCRB2 had an estimated 59% chance that adjusted OD would be higher for a positive than negative FS (95% CI: 51-67%). We performed one-sided Benjamini-Hochberg correction to control false discovery rate. Peptides from RESF1 and DTD2 survived at 5% (q=0.044; q=0.044; Figure 4E).

### A predictive model incorporating clinical variables shows good discrimination between SSA- SjD and combined control participants

We generated a regression model to predict SSA- SjD by incorporating IgG binding to our peptides into a model with clinical variables. After model selection, the predictive model included IgG binding to DTD2 (square-root transformed), unstimulated salivary flow (square-root), and ANA (other peptide binding and clinical factors did not add to the model; Figure 5A). This SjD prediction score discriminated between SSA- SjD and control participants. Area under the ROC curve (*C*-index) was 73.5% (95% CI: 66.0-79.9%), which decreased to 72.2% after correcting for optimism. Unstimulated salivary flow contributed the most to the model (single term deletion of unstimulated salivary flow yielded a more than 5.5 percentage point reduction in AUC) and second most important was binding to DTD2 (single term deletion of DTD2 binding yielded a more than 3.6 percentage point reduction in AUC). The model calculates a prediction score that is higher in SjD than combined control participants (Figure 5B). On cross validation, we showed that models using clinical predictors plus IgG binding to DTD2 had better overall prediction accuracy than models that used only the clinical variables (Figure 5C). Thus, inclusion of peptide binding improves the performance of models predicting SjD versus control.

**Figure 5.**
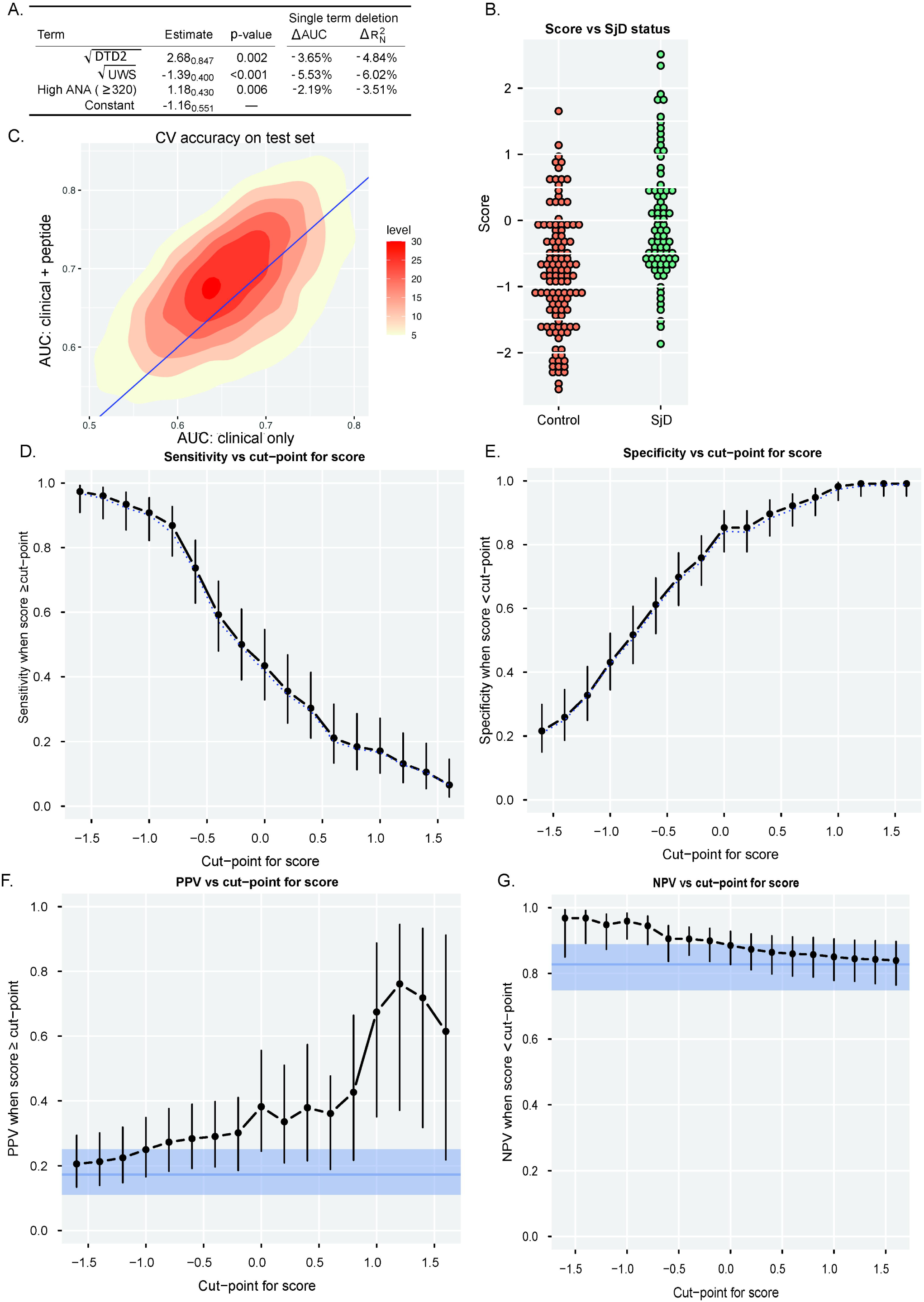
Models that incorporate binding to a peptide from DTD2 have good predictive ability for SjD. A) The selected predictive model incorporated three predictors (IgG binding to a peptide from DTD2, unstimulated salivary flow, and high ANA) with an AUC of 73.5% (95% CI: 66.0-79.9%), which decreased to 72.2% after correcting for optimism. The table shows estimated model coefficients and their standard errors as subscripts. The effects of single term deletion are shown; B) Dot plot showing the separation between SSA- SjD and combined controls by SjD prediction model score; C) In separate Monte Carlo cross validation, we repeatedly and randomly split the external validation data into 80% training and 20% testing; we used model selection on each training set, separately for clinical only variables or clinical plus peptide variables, and we built prediction rules on the test set. The improvement in AUC by including peptide variables is expressed in the shift above the diagonal line. Levels refer to histogram bin frequency in 10,000 training/test splits; D-E) Specificity and sensitivity graphed separately for cut-points of the score ranging from -1.6 to 1.6. Optimism-corrected values as dotted lines closely track the original values; F-G) Positive and negative predictive value graphed separately.

Sensitivity, specificity, positive predictive value, and negative predictive value are shown in Figure 5D-G. Using the selected predictive model, we can select thresholds that are either highly specific or highly sensitive, potentially confirming a SSA- SjD diagnosis without the need for biopsy in 7% of participants (n=5/76) or avoiding the need for a biopsy in 13% of controls that will not achieve a SjD diagnosis (n=15/116).

### A predictive model incorporating clinical variables shows good discrimination between FS positive and FS negative participants

We generated a regression model incorporating IgG binding to our peptides with clinical variables. The selected predictive model included IgG binding to DTD2 (square-root), unstimulated salivary flow (square-root), platelet count (log transformed), and ANA (Figure 6A). The *C*-index of the model was 71.6% (95% CI: 63.9-78.2%) and decreased to 69.3% after correcting for optimism. Binding to DTD2 contributed the most to the model (single term deletion of DTD2 yielded a more than 3.9 percentage point reduction in AUC) and the second most important was unstimulated salivary flow (single term deletion of unstimulated salivary flow yielded a 3.3 percentage point reduction in AUC). This final “FS prediction score” discriminated between FS positive and negative (Figure 6B). On cross validation, we showed that models using clinical predictors plus IgG binding to DTD2 had overall better prediction accuracy than models that used only the clinical variables (Figure 6C).

**Figure 6.**
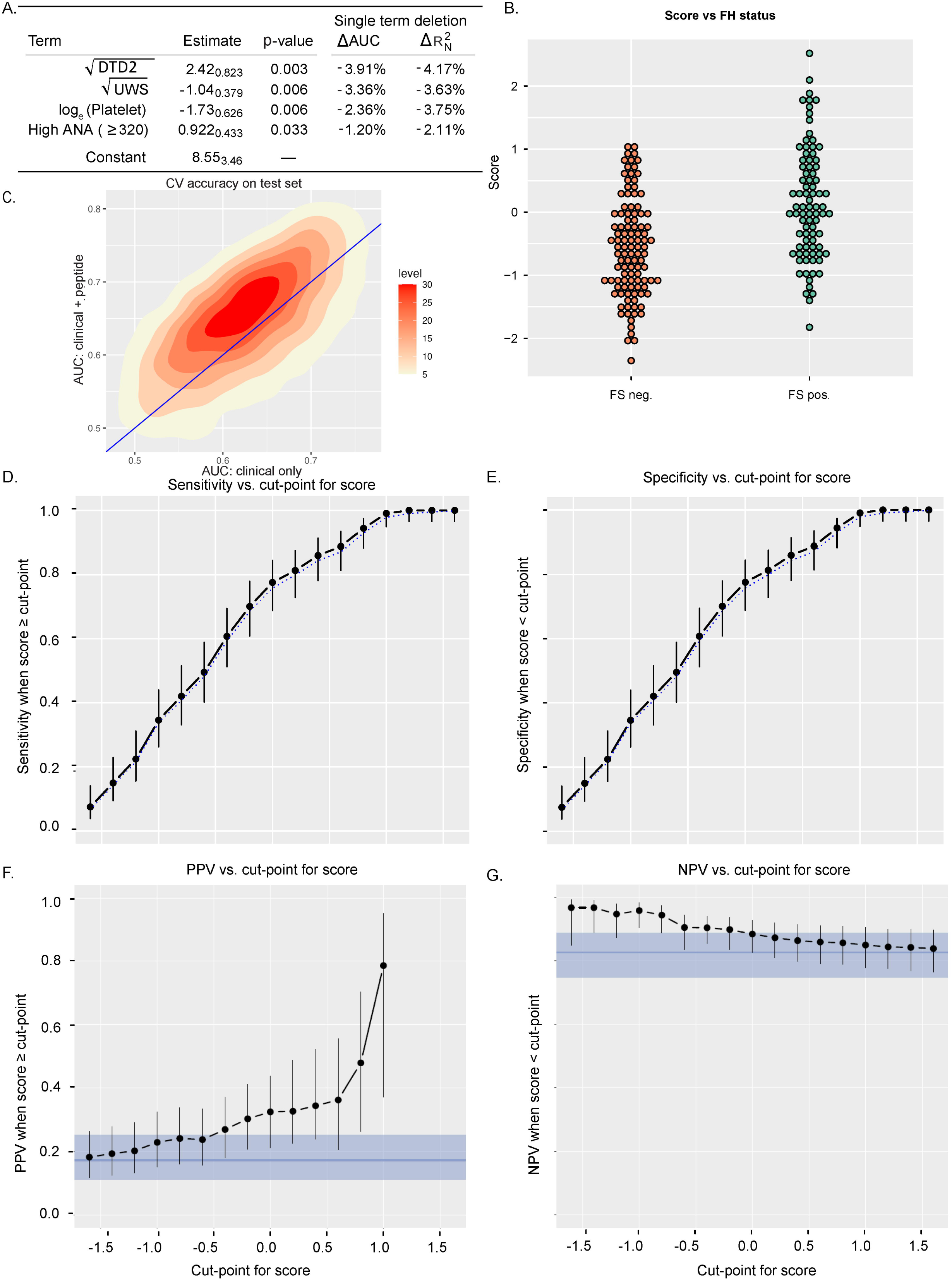
Models that incorporate binding to a peptide from DTD2 have good predictive ability for FS positivity. A) The selected predictive model incorporated four predictors (IgG binding to a peptide from DTD2, unstimulated salivary flow, platelet count, and high ANA) with an AUC of 71.6% (95% CI: 63.9-78.2%). The table shows estimated model coefficients and their standard errors in subscript. The effects of single term deletion are shown; B) Dot plot showing the separation of a model score between positive and negative FS groups; C) In separate Monte Carlo cross validation, we repeatedly and randomly split the external validation data into 80% training and 20% testing; we used model selection on each training set, separately for clinical only variables or clinical plus peptide variables, and we built prediction rules on the test set. The improvement in AUC by including peptide variables is expressed in the shift above the diagonal line. Levels refer to histogram bin frequency in 10,000 training/test splits; D-E) Specificity and sensitivity graphed separately for cut-points of the score ranging from -1.6 to 1.6. Optimism-corrected values as dotted lines and differ from original values by at most 2.6 or 1.8 percentage points for sensitivity and specificity, respectively; F-G) Positive and negative predictive value graphed separately.

We calculated sensitivity and specificity for FS prediction score cut-points (range -1.6 to 1.6; Figure 6D-E). Positive and negative likelihood ratios are shown (6F-G). Positive likelihood ratios could only be computed for cut-points ranging between -1.6 to 1.0, since none of the FS-positive group had calculated scores over 1.02. If we select a stringent positive score that indicates a biopsy will result in a positive FS (with no FS false positives), we could avoid the need for a salivary gland biopsy in 14% of patients (n=12/85). If we select a stringent negative score that indicates a salivary gland biopsy will be negative (no false negatives), we could avoid the need for a biopsy in 4% of patients (n=4/107).

## DISCUSSION

We describe new autoantibodies targeting peptides from DTD2 and RESF1 that are higher in SSA- SjD than relevant sicca and autoimmune controls. We also describe new autoantibodies targeting peptides from DTD2, RESF1, and SCRB2 that are higher in FS-positive than FS-negative participants. When DTD2 binding was combined with clinical features, we achieve good predictive discrimination between SSA- SjD and control participants and also between FS-positive compared to FS-negative participants.

Novel autoantibodies that help diagnose SSA- SjD fill a major gap in care for this patient population. The current standard for diagnosis of SSA- SjD patients requires a labial salivary gland biopsy. There are well recognized barriers to pursuing this biopsy including 1) concern from the patient about potential adverse effect, namely permanent numbness at the biopsy site; 2) finding a practitioner to perform the biopsy; and 3) identifying a pathologist with experience calculating a FS. Given these barriers, developing novel diagnostic tests using readily available sources is a major unmet clinical need. We showed that antibodies targeting peptides from DTD2 can be used along with standard clinical metrics to detect SSA- SjD or a positive FS with good discrimination. Indeed, binding to a peptide from DTD2 was the most important single term in our final model for FS prediction. Another benefit of these models was that we identified score cut points that will yield a high specificity or positive predictive value. Patients with a high SjD or FS prediction score might not need a labial salivary gland biopsy to confirm their diagnosis of SjD. On the other hand, we can select cut points with high sensitivity or negative predictive value. Patients with a very low score might not need to proceed to labial salivary gland biopsy because ultimately, they will not achieve criteria for SjD or have a positive FS.

Given the significant clinical need for novel diagnostic testing for SSA- SjD, others have also sought to detect autoantibodies. Farris et al. used a human proteome array with 19,500 proteins to identify 11 antibodies targeting novel proteins that were confirmed on a discovery and validation dataset [27]. Using a panel of 12 antigens, they developed a predictive model with an AUC of 0.88. Unfortunately, none of those protein target in the constructed model failed to predict SSA- SjD externally.

We found that autoantibodies in SSA- SjD bound a peptide from DTD2 most significantly. DTD2 recycles D-aminoacyl-tRNA to D-amino acids and free tRNA molecules and deacylates L-Ala [28]. Thus DTD2 might act as a proofreading mechanism; however, it has not been studied in more complex organisms, such as mice, where levels of DTD correlate with levels of neurotransmitters [29]. We also found SSA- SjD bound a peptide from RESF1 more than controls. RESF1 regulates gene expression and repressive epigenetic modifications. Specifically, it recruits SETDB1 for endogenous retrovirus silencing [30]. RESF1 also promotes embryonic stem cell self-renewal [31]. Finally, SCRB2 was bound more in FS positive than negative labial biopsies. SCRB2 is a lysosomal receptor for glucosylceramidase [32] and a receptor for enterovirus [33]. The absence of SCRB2 decreases macrophage and T-cell response in mouse models of crescentic glomerulonephritis [34] and Listeria infection [35]. It is unclear how IgG that recognizes linear epitopes in these proteins might contribute to SjD; indeed, it is not known if full-length proteins are bound by these autoantibodies. However, similar to other autoantigens in systemic autoimmunity (ex: Ro and histones), DTD2 and RESF1 interact with nucleic acids. Perhaps nucleic acids provide a danger signal via toll like receptors to stimulate the autoimmune response. A similar effect might be expected from enterovirus binding in the case of SCRB2. Further studies are needed to understand if and how these autoantibodies might be pathogenic.

We used innovative technology, a whole human peptidome array, for initial autoantibody identification. Motifs for the 469 peptides bound more in SSA- SjD than healthy controls on the array were associated with SjD-relevant proteins. For example, an identified motif is present in hnRNP. hnRNP is a pre-messenger RNA-binding protein that associates with RNA polymerase II transcripts to form protein-RNA complexes that act as substrates for RNA processing [36]. hnRNP is a recognized autoantigen in SjD [37] along with other systemic rheumatic diseases such as rheumatoid arthritis, systemic lupus erythematosus, and systemic sclerosis, among others [38–40]. Another motif is present in complement c1q tumor necrosis factor-related protein, CTRP2. CTRP2 is a protein secreted in tissues such as adipose, lung, liver, testes, and uterus. It regulates insulin tolerance and lipolytic enzymes [41]. CTRP2 is similar in structure to adiponectin in the globular domain and can induce phosphorylation of AMP-activated protein kinase (AMPK) and Akt [42]. AMPK activation is salient to SjD because it inhibits mTOR which is implicated in cell growth, survival and proliferation and also regulates T cell differentiation [43]. We and others have shown evidence that metformin, through inhibition of mTOR, might improve SjD [44, 45]. Thus, motifs bound in SSA- SjD might provide some insight into the functional relevance of our antigenic targets.

We also found interesting GO themes like cell junction and postsynaptic signaling that included genes such as glutamate receptor ionotropic (NMDA 3A), Cytoplasmic polyadenylation element-binding protein 4 (CPEB4), and TANC1, among others. Glutamate receptor antibodies are described previously in SjD and are responsible for decreased receptor expression, impaired signaling, and neuronal damage [46, 47]. CPEB4 regulates the unfolded protein response and is required for cytokinesis [48]; it is required for resolution of induced macrophage inflammatory response [49]. CPEB might relevant to SjD because it stabilizes mRNA that encode for negative feedback regulators of inflammatory response, such as LPS [49].

Strengths of our study include the innovative whole human peptidome array and our novel statistical approach to identify new peptide targets. We include clinically relevant controls that mimic the population who would be referred for a salivary gland biopsy or a possible SjD diagnosis. Finally, we use robust sample sizes to validate our array findings in a large independent population. Limitations of this study include the linear formation of the array and ELISA peptides, which do not have the conformational structure of native proteins. The peptide array does not account for native protein modifications, such as glycosylation.

In conclusion, we present novel autoantibodies in SSA- SjD compared to autoimmune- and sicca-controls that can be used to predict an SjD diagnosis or abnormal FS on labial salivary gland biopsy with good predictive value.

## Data Availability

all data produced in the present study are availbe upon reasonable request to the authors

## ACKNOWLEDGEMENTS

We thank Jacques Galipeau MD for his mentorship and support.

